# Cardiology Knowledge Assessment of Retrieval-Augmented Open versus Proprietary Large Language Models

**DOI:** 10.1101/2025.09.11.25335607

**Authors:** Constantine Tarabanis, Shaan Khurshid, Areti Karamanou, Rodo Piperaki, Lucas A. Mavromatis, Aris Hatzimemos, Dimitrios Tachmatzidis, Constantinos Bakogiannis, Vassilios Vassilikos, Patrick T. Ellinor, Lior Jankelson, Evangelos Kalampokis

## Abstract

**Objectives:** To evaluate the performance of open and proprietary LLMs, with and without Retrieval-Augmented Generation (RAG), on cardiology board-style questions and benchmark them against the human average.

**Materials and Methods:** We tested 14 LLMs (6 open-weight, 8 proprietary) on 449 multiple-choice questions from the American College of Cardiology Self-Assessment Program (ACCSAP). Accuracy was measured as percent correct. RAG was implemented using a knowledge base of 123 guideline and textbook documents.

**Results:** The open-weight model DeepSeek R1 achieved the highest accuracy at 86.7% (95% CI: 83.7–89.9%), outperforming proprietary models and the human average of 78%. GPT 4o (80.8%, 95% CI: 77.2–84.5%) and the commercial platform OpenEvidence (80.4%, 95% CI: 76.7–84.0%) demonstrated similar performance. A positive correlation between model size and performance was observed within model families, but across families, substantial variability persisted among models with similar parameter counts. After RAG, all models improved, and open-weight models like Mistral Large 2 (78.0%, 95% CI: 74.1–81.8) performed comparably to proprietary alternatives like GPT 4o.

**Discussion:** Large language models (LLMs) are increasingly integrated into clinical workflows, yet their performance in cardiovascular medicine remains insufficiently evaluated. Open-weight models can match or exceed proprietary systems in cardiovascular knowledge, with RAG particularly beneficial for smaller models. Given their transparency, configurability, and potential for local deployment, open-weight models, strategically augmented, represent viable, lower-cost alternatives for clinical applications.

**Conclusion:** Open-weight LLMs demonstrate competency in cardiovascular medicine comparable to or exceeding that of proprietary models, with and without RAG depending on the model.

**Author Summary:** In this work, we set out to understand how today’s artificial intelligence systems perform when tested on the kind of questions cardiologists face during board examinations. We compared a wide range of large language models, including both freely available “open” models and commercial “proprietary” ones, and also tested whether giving the models access to trusted cardiology textbooks and guidelines could improve their answers. We found that the best open model actually outperformed all of the commercial models we tested, even exceeding the average score of practicing cardiologists. When we gave the models access to medical reference material, nearly all of them improved, with the biggest gains seen in the smaller and weaker models. This shows that careful design and support can allow smaller, more accessible systems to reach high levels of accuracy. Our results suggest that open models, which can be used locally without sending sensitive patient information to outside servers, may be a safe and cost-effective alternative to commercial products. This matters because it could make powerful AI tools more widely available across hospitals and clinics, while also reducing risks related to privacy, transparency, and cost.

## Introduction

Large language models (LLMs) have been proposed to hold transformative potential in cardiovascular medicine[1,2], with applications ranging from automated note generation to clinical consultations[3]. To optimize their performance in medical contexts, various techniques have been developed, including domain-specific fine-tuning and retrieval-based methods[4]. An example of the latter is Retrieval-Augmented Generation (RAG), which aims to improve performance by incorporating relevant content from a provided document set during response generation[4]. However, before these models can be reliably applied in clinical practice, they should undergo rigorous assessment. One such initial evaluative step has involved the investigation of LLM performance on the United States Medical Licensing Examination[5,6] and board exams in internal medicine[7], ophthalmology[8] and other fields[9–11]. The variability in model accuracy across internal medicine subspecialties[7] underscores the importance of domain-specific evaluations. Such analyses are lacking in cardiology and would serve as a first step to benchmarking LLMs’ fund of knowledge against that of practicing cardiologists.

In this context, comparisons between open and proprietary LLMs are needed, as the former offer enhanced patient data safety, improved bias evaluation, and greater customization potential[12]. Additionally, analyzing the relationship between model size and exam performance is key to estimating the computational resources required for factually accurate medical LLMs. Understanding how model characteristics and retrieval-based techniques impact accuracy is essential for ensuring real-world feasibility. Yet, these relationships remain minimally explored in the cardiovascular literature. In this study, we evaluate open and proprietary LLMs of varying model parameter size, with and without the application of RAG, on cardiology board exam-style questions from the American College of Cardiology Self-Assessment Program (ACCSAP).

## Results

A total of eight proprietary and six open LLMs (**Supplementary Table 2**) were evaluated on 449 cardiology board-style questions spanning a broad range of specialty-specific subject areas (**Supplementary Table 1**). The models’ performance, expressed as a percentage of correct answer choices, is summarized in **Figure 1A**. The top five performing models in descending order include: DeepSeek R1 (86.7%, 95% CI: 83.7-89.9%), GPT 4o (80.8%, 95% CI: 77.2-84.5%), Claude 3.7 Sonnet (76.6%, 95% CI: 72.7-80.5%), GPT 4 Turbo (73.7%, 95% CI: 69.6-77.9%), and Mistral Large 2 (73.7%, 95% CI: 69.6-77.8%). The two highest performing open models included DeepSeek R1 and Mistral Large 2 with 671 and 123 billion parameters, respectively. We also assessed OpenEvidence, an LLM-powered proprietary commercial platform designed to deliver clinical answers to healthcare professionals, which achieved a performance of 80.4% (95% CI: 76.7-84.0%) on par with the second highest performing model, GPT 4o. Interestingly, Llama 3.1 8B exhibited a bias towards response “A” resulting in its observed underperformance. DeepSeek was the only LLM to outperform the human average of 78%, while the 95% confidence interval of GPT 4o encompassed the human user-derived mean. Focusing on one top performing LLM per model family, **Figure 1B** depicts model accuracy across key cardiology subject areas. In this context, DeepSeek R1 achieved equivalent or superior performance in 7 of 11 subject areas, followed by Claude 3.7 Sonnet in 3 of 11, and GPT-4o in one.

**Figure 1.**
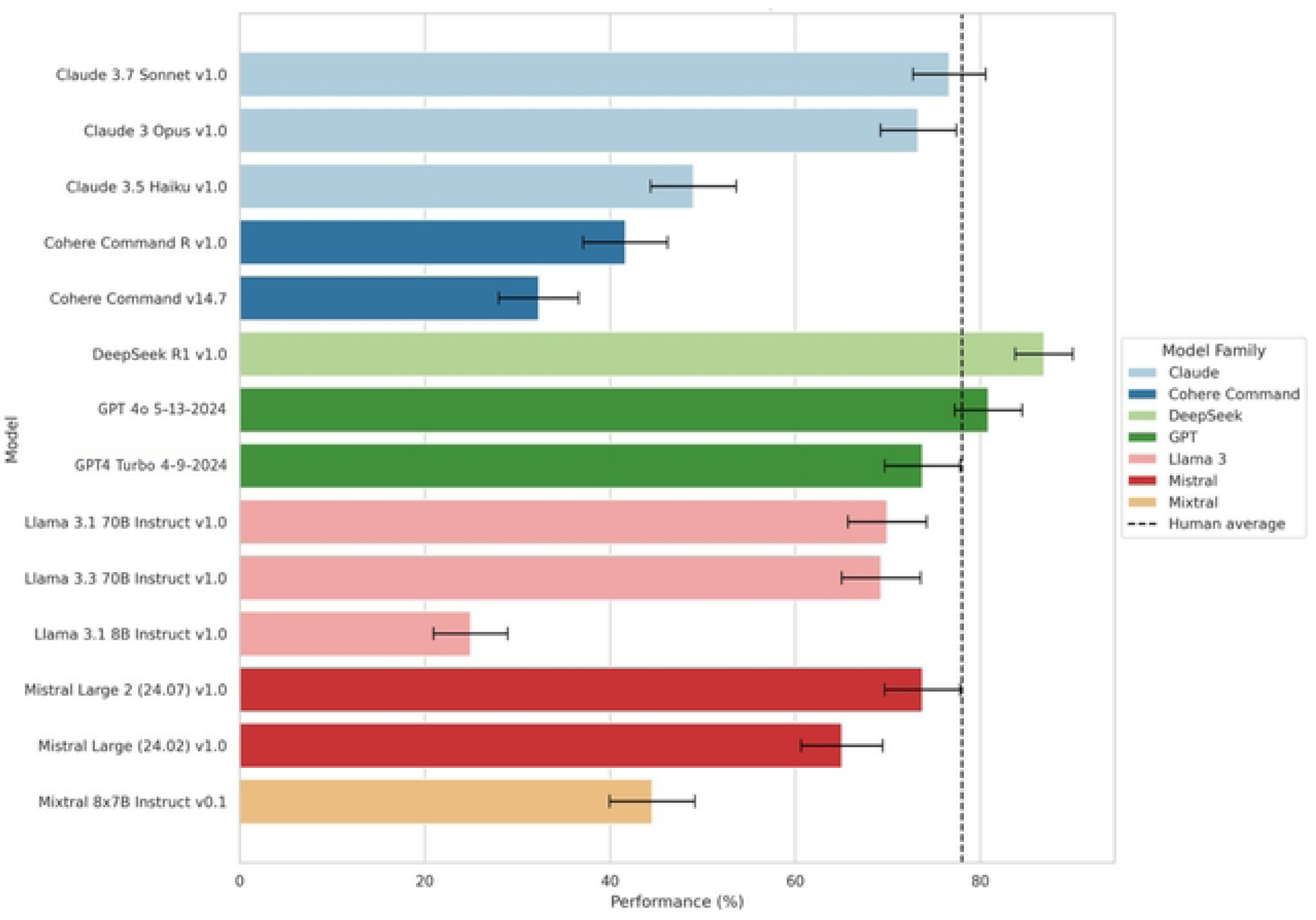
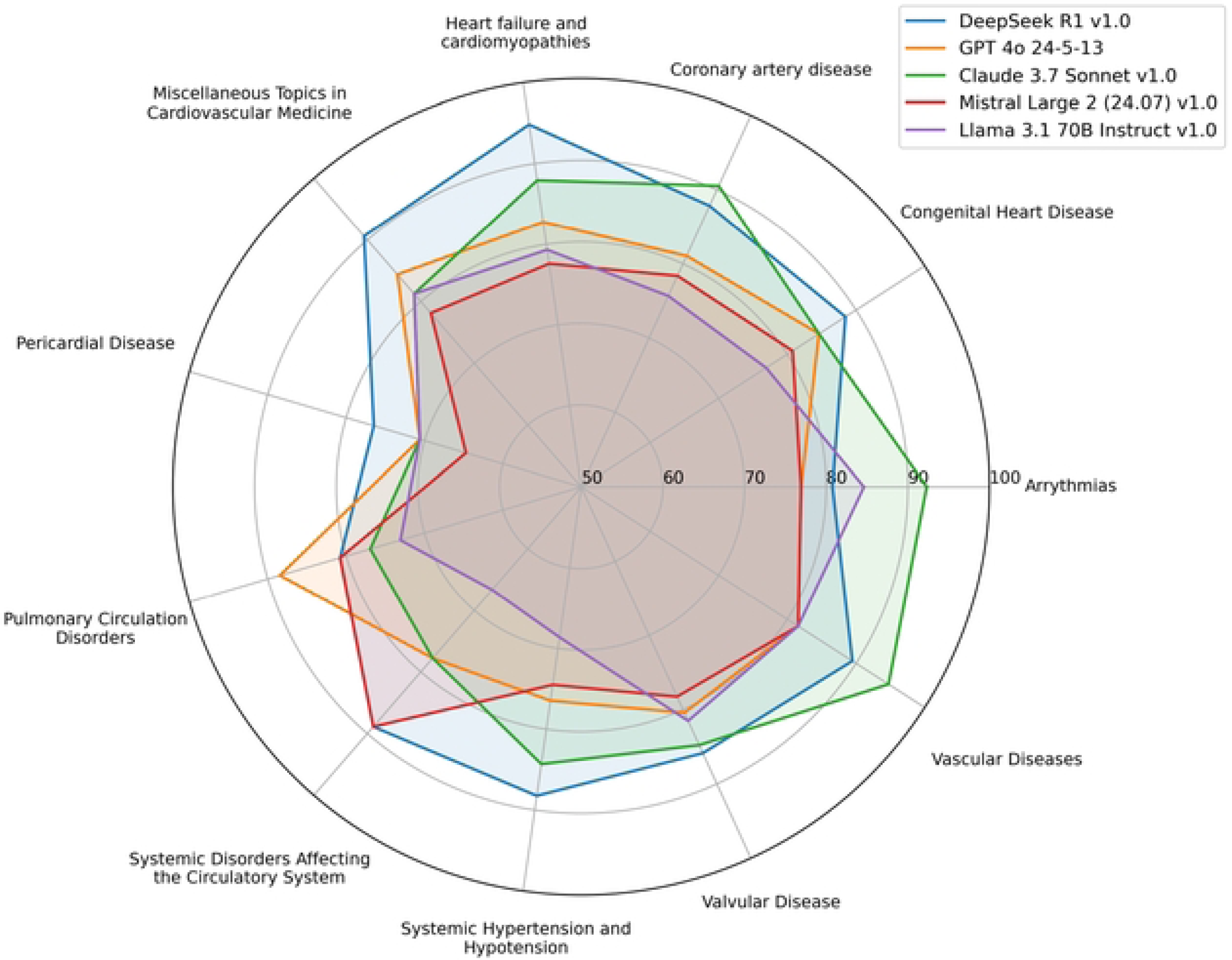
**(A)** Performance comparison of various LLMs across different model families on cardiology board-style questions, with the mean human performance indicated by a dashed line. **(B)** Radar chart displaying the percentage of correctly answered cardiology board-style questions by the top-performing LLM from each model family, categorized across major cardiology thematic areas.

The highest performing model, DeepSeek R1, exhibited the highest pairwise agreement scores with the other top four performing models, both open and proprietary (**Figure 2A**). The pairwise agreement scores assessed concordance for each question and the highest score achieved was 0.76 between Mistral Large 2 (open) and GPT 4o (proprietary). All remaining scores were lower, indicating a higher degree of variability in the answers each model selected. The percentage of correct answers for each model was graphed against the log-transformed number of model parameters (**Figure 2B**). The observed positive correlation suggests that both across and within model families, increasing the number of model parameters results in improved exam performance (**Figure 2B**).

**Figure 2.**
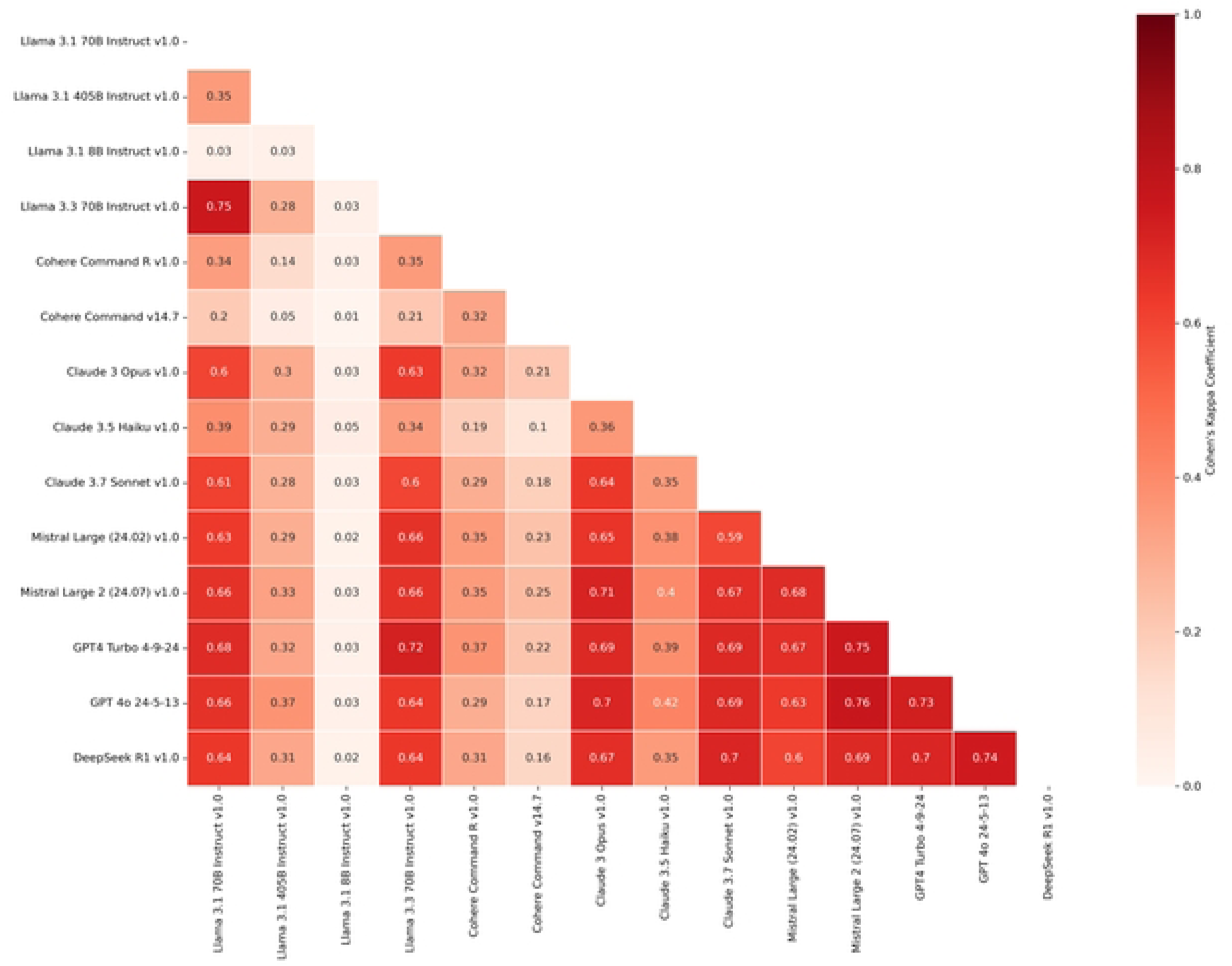
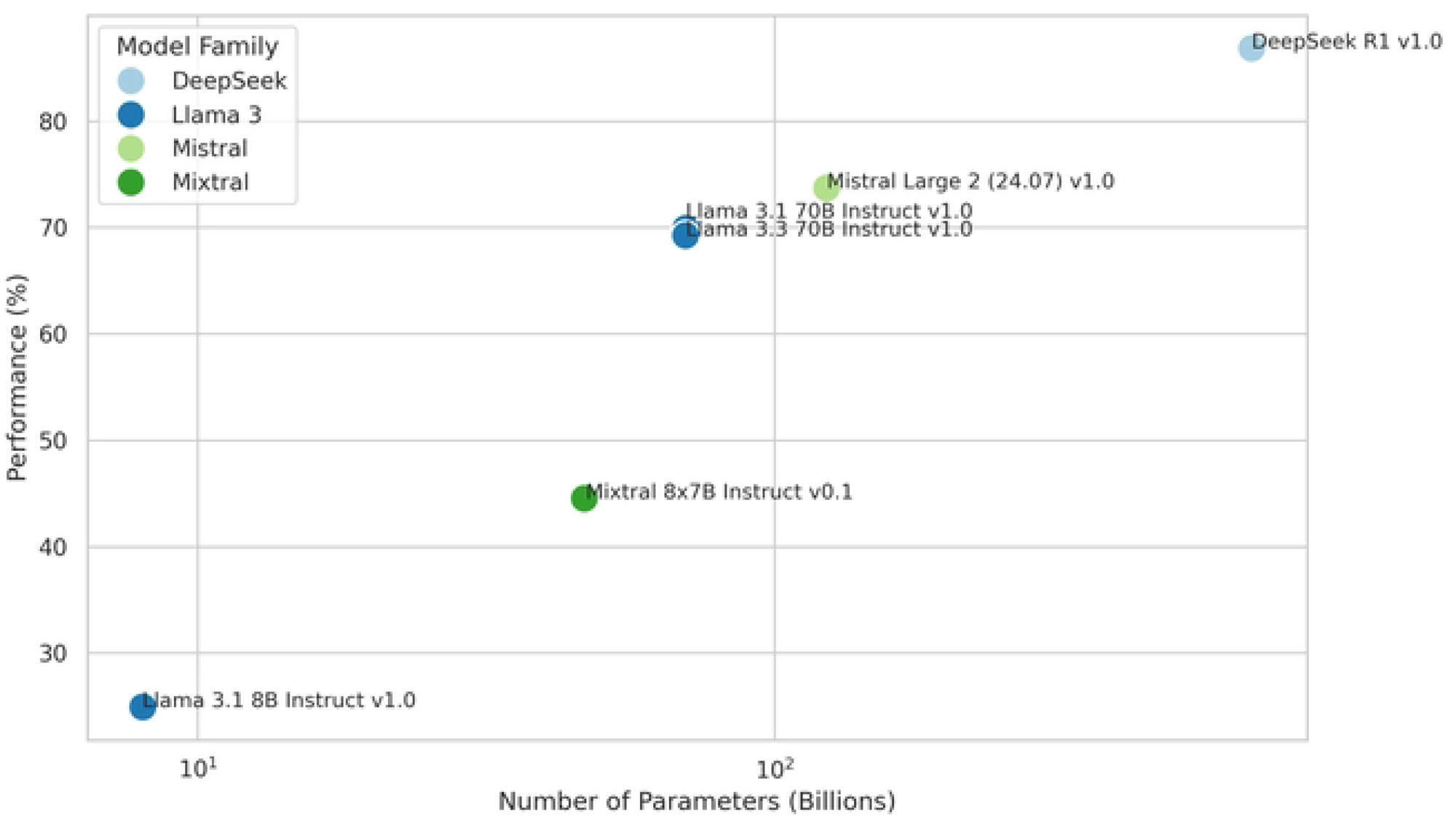
**(A)** Heatmap showing the pairwise Cohen’s Kappa coefficients for the answer choices of different LLMs on cardiology board-style questions, with red indicating stronger agreement between models. **(B)** Scatter plot showing the relationship between the log-transformed number of model parameters (in billions) and response accuracy (%) on cardiology board-style questions across different LLMs.

Comparison of model performance before and after integrating RAG (**Figure 3**) illustrates that all models showed improvement, though to varying degrees. After RAG, the top five performing models in descending order are: DeepSeek R1, Claude 3.7 Sonnet, GPT 4o, Claude 3 Opus and Mistral Large 2. Hence the Claude models 3.7 Sonnet and 3 Opus overtook GPT 4o and GPT4 Turbo, respectively. DeepSeek R1 achieved the smallest absolute (0.8%) and relative improvement with RAG, yet remained the top performing model achieving 87.5% (95% CI: 84.5-90.6%). Among the top five performing models, Claude 3.7 Sonnet and Claude 3 Opus achieved the greatest relative performance improvements at 12.2% and 13.3%, resulting in overall accuracies of 86.0% (95% CI: 82.8-89.2%) and 83.1% (95% CI: 79.6-86.5%), respectively. GPT 4o and Mistral Large 2 exhibited similar relative performance improvements of 5% and 5.7% achieving overall accuracies of 84.9% (95% CI: 81.5-88.2%) and 78.0% (95% CI: 74.1-81.8%), respectively. Overall, the greatest performance gains were observed in models that initially performed the worst prior to RAG integration. Llama 3.1 8B improved by 130% (57.5, 95% CI: 52.9-62.0%), Cohere Command v14.7 by 56.6% (50.6%, 95% CI: 45.9-55.2%), and Claude 3.5 Haiku by 48.2% (72.6, 95% CI: 68.5-76.7%). Interestingly, the RAG process resolved Llama 3.1 8B’s bias issue towards “A” responses.

**Figure 3.**
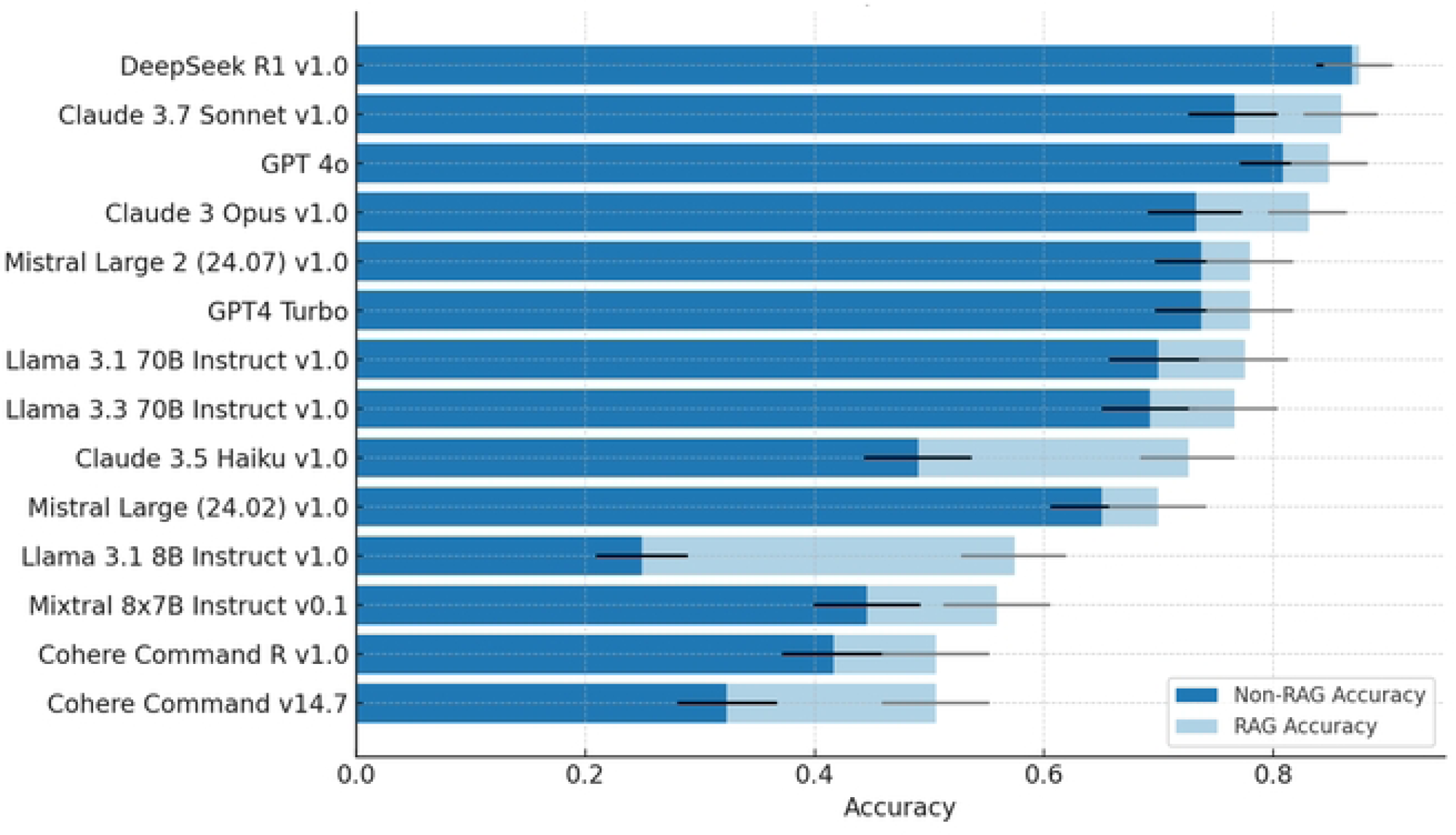
Performance comparison of various LLMs before and after applying RAG in a descending order of accuracy.

## Discussion

In this study, we systematically compared the accuracy of 14 LLMs from various families, across varying model sizes and degrees of transparency (open versus proprietary), on 449 cardiology board exam-style questions (**Central Illustration**). The recently developed open-weight model DeepSeek R1 outperformed all proprietary alternatives (including OpenAI’s GPT 4o), led across nearly all cardiology subdomains, and was the only model to exceed the human test-taking average without RAG. While a positive correlation between model size and accuracy was observed within model families, performance varied significantly across families, even among LLMs with similar parameter counts. The use of RAG improved performance across all models, with open-weight models achieving performance levels comparable to proprietary ones.

LLMs are already being incorporated into medical practice[13], despite the lack of an established, systematic evaluation protocol prior to clinical use. A recent publication proposed the development of Artificial Intelligence Structured Clinical Examinations (AI-SCEs)[14]. Emulating Objective Structured Clinical Examinations (OSCEs) used in medical trainees’ assessments, AI-SCEs would involve complex real-world clinical scenarios designed by interdisciplinary teams of clinicians, computer scientists, and medical researchers[14]. Until formal protocols are established, however, medical LLMs should at least demonstrate high factual accuracy on the same medical examinations used to certify physicians[15]. Although this alone is insufficient to establish clinical utility, it is a key first evaluative step. This way, LLM performance can be benchmarked against the existing knowledge base of practicing clinicians. The current work therefore addresses a key gap in the current understanding of LLM competency in cardiovascular medicine.

The leading performance of DeepSeek challenges the prevailing assumption that proprietary models inherently outperform their open counterparts, instead suggesting that large open models may already be capable of rivaling the most advanced commercial systems even in specialized medical knowledge. This is particularly important for the medical field, because open LLMs offer key advantages over proprietary ones. Proprietary LLMs often necessitate uploading private data to online servers and the lack of transparency raises questions about how protected patient information might be stored or used, such as for further training purposes[16]. In contrast, open LLMs can operate offline on secure platforms protecting patient confidentiality by reducing the likelihood of data breaches[12]. Additionally, the open-weight structure of LLMs, like DeepSeek and Mistral Large 2, renders them customizable to specific healthcare needs and may facilitate more comprehensive bias evaluation[12].

Despite being positioned as a specialized clinical platform for evidence-based answers, OpenEvidence achieved a score of 80.4%, similar to GPT 4o and lower than DeepSeek R1, both general-purpose LLMs not developed for medical use, with the latter notably being open-weight. This raises important concerns about the added value of proprietary platforms that are already being used in clinical practice and often lack independent external validation. In cardiovascular medicine, this limited assessment suggests that LLM-powered commercial platforms are not necessarily superior to freely accessible open-weight models such as Mistral Large 2 post-RAG or DeepSeek R1. This raises broader questions about whether the restricted access, lack of transparency, and commercial cost associated with these platforms are justified.

Additionally, we demonstrate a trend within LLM families (e.g., Mistral and Llama), where performance in cardiology exam questions improves as parameter size increases. This suggests that, within a single family, larger models generally have better factual accuracy in accordance with prior results[15]. This has important implications for the use of these models in real-world healthcare settings, since the larger the model the greater the computational resource demands. This could limit the accessibility of highly accurate LLMs to smaller healthcare systems, pushing them toward reliance on commercial LLM products or partnerships with larger health systems that have the necessary computational infrastructure. These dynamics risk exacerbating existing disparities in healthcare quality, disproportionately affecting vulnerable populations. Less resourced, more rural, and relatively marginalized healthcare systems may face greater barriers to accessing these advanced tools, further entrenching inequities[17].

At the same time, we also observed considerable differences in performance across different LLM families independent of varying parameter sizes. For instance, Llama 3.1 and 3.3 70b had a comparable (overlapping 95% CIs) performance to Mistral Large 2, despite the latter having an additional 53 billion parameters. This discrepancy indicates that factors beyond the sheer size of a model, such as differences in model architecture, the quality and diversity of training data, and pre-training objectives, have a significant impact on performance. Coupled with our observation that the lowest-performing models (often also the smallest by parameter count) demonstrated the greatest relative improvements after RAG integration, this suggests that smaller, open-weight models can be strategically augmented to close the performance gap. Such gains may also be achievable by focusing fine-tuning efforts on areas of relative deficiency, as identified in our analysis of variability across cardiology subdomains. With targeted retrieval or fine-tuning, these models may achieve accuracy comparable to much larger counterparts without incurring the computational demands or privacy risks associated with proprietary large-scale LLMs[18]. Future work is warranted to define the optimal tradeoff of model size (and subsequent resource-use) versus performance, which may vary according to specific intended applications.

The present study’s limitations include the aforementioned inability to test ABIM questions directly and the use of solely text-based questions. This exclusion of image-based questions reflects a constraint of LLMs in their current form and could result in the disproportionate representation of only certain subject areas. Additionally, the absence of a definitive gold-standard measure of human performance is another key limitation. While the rolling average of approximately 78% from ACCSAP provides a useful benchmark, it is significantly influenced by factors such as repeated question exposure and evolving user performance over time.

## Conclusion

This study demonstrates that both open and proprietary LLMs can achieve high performance on cardiology board exam-style questions, with the open-weight model DeepSeek R1 outperforming all proprietary alternatives and exceeding the human test-taking benchmark. While model size generally correlated with accuracy within families, cross-family comparisons revealed that parameter count alone does not determine performance, underscoring the influence of model architecture and design. Importantly, RAG substantially boosted the performance of smaller, lower-performing models, and in some cases corrected systemic biases. These findings suggest a promising path forward: smaller, open-weight models, when strategically augmented with retrieval or fine-tuned based on subspecialty-specific deficiencies, may achieve high accuracy without the privacy risks or infrastructure demands of large proprietary systems.

## Materials and Methods

### Question selection

Since the American Board of Internal Medicine (ABIM) does not have any publicly available testing material, we used ACCSAP questions that are similar in complexity to the multiple-choice questions from the ABIM cardiovascular disease certification exam. ACCSAP questions cover a wide range of knowledge from diagnosis to medical management and are commonly used to prepare for the ABIM certification exam. ACCSAP reports the average percentage of correct responses by human users for each question. This human test-taking average can change over time, reflecting the cumulative performance of all user responses per question within the most recent iteration of the question bank. As a proprietary database existing behind a paywall, ACCSAP questions are not freely accessible on the internet and should be outside the scope of LLM training data. This was confirmed by a Google index search using the exact first ten words of each question prompt, with the date filter set to before March 15, 2025. All questions containing clinical images were excluded and 449 questions were randomly selected comprising all major subject areas as defined by the ACCSAP, with their percentage distribution included in **Supplementary Table 1**. Ethical approval was waived for this study, as it did not involve human participants or identifiable patient data.

### Large language model selection

We evaluated a number of contemporary LLMs, which are listed in **Supplementary Table 2** along with their access type, release date, context length, and parameter size (when known). LLM openness refers to the degree to which a model’s components (e.g., architecture, pretrained weights, training code, training datasets) are publicly available under licenses that allow independent use, modification, and sharing. A proprietary model (Claude, GPT, Mistral Large 24.02 v1.0) restricts access to its components, offering use only through an Application Programming Interface (API) or licensed platforms without allowing inspection, modification, or self-hosting. An open weight model (DeepSeek, Llama 3.1/3.3, Mistral Large 24.07 v1.0) publicly releases its pretrained weights for download and local use, but other model components, such as training data/code and data curation methods, are typically withheld, limiting full reproducibility. An open-source model makes all key components publicly available under an open-source license, allowing anyone to use, modify, reproduce, and distribute the model without restriction. Based on the aforementioned definitions, the present study evaluated proprietary and open weight models, with the latter referred to as “open” hereafter.

### Inference and Hyperparameter Settings

Inferences from each model were obtained via SageMaker or Bedrock Amazon Web Services (AWS, Seattle, WA, USA). The LLMs, according to the extent to which each of them could be parameterized, were configured for temperature, top-P and output token limit. Temperature is the parameter responsible for the degree of smoothing of the output token probability distribution. As temperature rises, the probability distribution approximates the uniform distribution, effectively reducing the output token selection to random choice. Top-P, on the other hand, sets a cutoff according to the cumulative probability distribution of the tokens, limiting the tokens to be considered for selection. All experiments were conducted with a temperature of 0, a top-P of 0.9 and a max output token limit of 512[19]. This combination of hyperparameters was selected to ensure high reproducibility (low temperature), while leaving room for creative expression (high top-P). Setting the temperature to zero ensures deterministic and fully reproducible outputs by always selecting the highest-probability token at each step. The top-P parameter of 0.9 restricts the token selection to the most probable tokens that cumulatively account for 90% of the probability mass, enhancing the coherence and relevance of the generated text[19].

### Performance evaluation

All special characters except for punctuation marks were removed from each ACCSAP question prior to model inference. To ensure that models provided a clear, single answer choice along with a supporting explanation, each question was wrapped into the following prompt: “You are taking an examination that assesses your knowledge in cardiology within the field of medicine. For each question, you answer first with the letter corresponding to the correct answer and then the explanation as to why you have selected the answer. Question: {dataset question}”. An exception was made for the OpenAI models, where the standardizing prompt was used as the system prompt, while the question was passed as the user’s query. After inference, model outputs were reviewed to identify the selected answer choice for each question and calculate each model’s test taking accuracy.

### Retrieval-Augmented Generation

Retrieval-Augmented Generation (RAG) is a technique that attempts to improve language model performance by retrieving and incorporating relevant content from an external, provided document set during the response generation process. The impact of RAG on the aforementioned model performance was investigated. The entire RAG process for all models was based on Amazon’s Bedrock Knowledge Base service. A total of 123 documents were used, including 14 textbooks (primarily companions to *Braunwald’s Heart Disease*) and 109 cardiology guidelines and expert consensus documents from the American College of Cardiology and European Society of Cardiology from 2012 to 2024 (**Supplementary Table 3**). The documents were divided into smaller segments (chunks) following a fixed-size chunking approach. The maximum number of tokens allowed per chunk during document segmentation was set to 500 with an overlap percentage of 20%. The resulted chunks were converted into a vector representation through the amazon.titan-embed-text-v1 embedding model and stored in Amazon OpenSearch. In order to improve the retrieval of relevant chunks, each multiple-choice question was reformulated by each LLM based on the orchestration prompt template (**Supplementary Figure 1**). The reformulated question was then transformed into a vector representation using the same embedding model and compared to all chunks following a dynamically adjusted approach (hybrid/semantic). The maximum number of retrieved chunks was set to 15. These chunks where then provided to the generation prompt template (**Supplementary Figure 2**), which produced the final prompt that was submitted to the LLM in order to produce the final response.

## Data Availability

N/A ACCSAP questions are under copyright. Code already available in supplemental material and can provide additional code upon request.

## Acknowledgments

AWS resources were provided by the National Infrastructures for Research and Technology GRNET and funded by the EU Recovery and Resiliency Facility. Dr. Ellinor is supported by grants from the National Institutes of Health (RO1HL092577, RO1HL157635, R01HL177209), from the American Heart Association (961045), from the European Union (MAESTRIA 965286) and from the Fondation Leducq (24CVD01).

